# Health and Economic Consequences of Universal Paid Sick Leave Policies During the COVID-19 Pandemic

**DOI:** 10.1101/2022.01.13.21268270

**Authors:** David Naimark, Juan David Rios, Sharmistha Mishra, Beate Sander, Petros Pechlivanoglou

## Abstract

**Importance:** Universal paid sick-leave (PSL) policies have been implemented in jurisdictions to mitigate the spread of SARS-CoV-2. However empirical data regarding health and economic consequences of PSL policies is scarce.

**Objective:** To estimate effects of a universal PSL policy in Ontario, Canada’s most populous province.

**Design:** An agent-based model (ABM) to simulate SARS-CoV-2 transmission informed by data from Statistics Canada, health administrative sources, and from the literature.

**Setting:** Ontario from January 1^st^ to May 1^st^, 2021.

**Participants:** A synthetic population (1 million) with occupation and household characteristics representative of Ontario residents (14.5 million).

**Exposure:** A base case of existing employer-based PSL alone versus the addition of a 3-or 10-day universal PSL policy to facilitate testing and self-isolation among workers infected with SARS-CoV-2 themselves or because of infected household members.

**Main Outcome(s) and Measure(s):** Number of SARS-CoV-2 infections and COVID-19 hospitalizations, worker productivity, lost wages, and presenteeism (going to a workplace while infected).

**Results:** If a 3- and 10-day universal PSL were implemented over the 4-month study period, then compared with the base-case, the PSL policies were estimated to reduce cumulative SARS-CoV-2 cases by 85,531 (95% credible interval, CrI -2,484; 195,318) and 215,302 (81,500; 413,742), COVID-19 hospital admissions by 1,307 (-201; 3,205) and 3,352 (1,223; 6,528), numbers of workers forgoing wages by 558 (-327;1,608) and 7,406 (6,764; 8,072), and numbers of workers engaged in presenteeism by 24,499 (216; 54,170) and 279,863 (262,696; 295,449). Hours of productivity loss were estimated to be 10,854,379 (10,212,304; 11,465,635) in the base case, 17,446,525 (15,934,321; 18,854,683) in the 3-day scenario, and 26,127,165 (20,047,239; 29,875,161) in the 10-day scenario. Lost wages were $5,256,316 ($4,077,280; $6,804,983) and $12,610,962 ($11,463,128; $13,724,664) lower in the 3 day and 10 day scenarios respectively, relative to the base case.

**Conclusions and Relevance:** Expanded access to PSL is estimated to reduce total numbers of COVID-19 cases, reduce presenteeism of workers with SARS-CoV-2 at workplaces, and mitigate wage loss experienced by workers.

**Competing interests:** The authors have no competing interests relevant to this article to disclose.

**Funding:** Supported by COVID-19 Rapid Research Funding (C-291-2431272-SANDER). This research was further supported, in part, by a Canada Research Chair in Economics of Infectious Diseases held by Beate Sander (CRC-950-232429). The study sponsor had no role in the design, collection, analysis, interpretation of the data, manuscript preparation or the decision to submit for publication.

**Author Contributions:** Conceptualization: PP, JDR, BS, DN

Data Curation: PP, JDR, BS, DN

Formal Analysis: PP, JDR, DN

Methodology: PP, JDR, BS, DN

Supervision: PP, DN, BS

Validation: PP, JDR, BS, DN

First Draft: PP, JDR, BS, DN

Review and Edit

PP, JDR, BS, DN

**Key points:** *Question:* What could be the health and economic consequence of more generous paid sick leave policies in the context of the COVID-19 pandemic?

*Findings:* More generous policies are estimated to reduce SARS-CoV-2 infections (and thus COVID-19 hospitalizations), lost wages and presence of individuals with infection at workplaces.

*Meaning:* More generous paid sick leave can be a valuable addition to other COVID-19 public health interventions.

## Introduction

COVID-19-related paid sick leave (PSL) policies, implemented or expanded in many OECD countries allow workers to isolate without forgoing wages.^1–3^ In the United States, states with expanded PSL policies had reduced rates of SARS-CoV-2 infections compared to states with limited PSL policies.^4^ Pre-pandemic, 60% of workers lack employer-provided PSL in Ontario, Canada,^2^ and rely on a federally administered program offering income support days to weeks *after* confirmed SARS-CoV-2. This results in a reduction and disruption of wages if workers without PSL self-isolate, take time off to undergo testing, or quarantine. ^2^ We estimated the health and economic consequences of universal PSL policies in Ontario for workers without employer-provided PSL.

## Methods

An agent-based model (ABM) of SARS-CoV-2 transmission simulated a synthetic population of 1 million individuals representative of the Ontario population (14.5 million) employing data from Statistics Canada’s Social Policy Simulation Database/Model.^5^ This ABM has been used to estimate the spread of SARS-CoV-2 in schools (**Supplementary Methods: Text 1)**.^6^ The ABM was modified by assigning simulated workers to employer-based PSL depending on age, sex, marital status, family income, and occupational classification employing data from Statistics Canada’s General Society Survey (**Supplementary Methods: Text 2)**.^7^ To estimate lost wages, hours worked and hourly wages were assigned to workers based on occupational classification and full-time/part-time status. ^8,9^

A four-month period (January 1 to May 1, 2021) was simulated representing the third COVID-19 wave in Ontario. Health outcomes included the number of SARS-CoV-2 infections and COVID-19 hospitalizations. The mechanism underlying the health benefits of expanded PSL was modelled as reduced contacts in workplaces. Economic outcomes included estimated productivity loss (reduction in hours worked), lost wages and presenteeism (going to work with detected or symptomatic infection). Model outcomes were scaled by 14.5 to generate Ontario-level estimates.

Three scenarios were modelled: 1) *<u>base case</u>* where only employer-based PSL was available, 2) *<u>3-day universal PSL</u>* where the latter workers continued to receive employer-based PSL and, in addition, all remaining workers were eligible for three PSL days, and similarly 3) *<u>10-day universal PSL</u>* where remaining workers were eligible for ten PSL days (model assumptions are provided in **Supplementary Text 3**). A key assumption in the model is that workers would take advantage of PSL if available, to self-isolate or quarantine and, without PSL, they would be unable to take these steps and would engage in presenteeism, unless severely ill. Similarly, for working household members, who were unable to work remotely and had a household member infected and detected/symptomatic with SARS-CoV-2, the ability to self-isolate or quarantine depended on access to PSL. We also modeled the economic consequences of PSL on work hours lost due to SARS-CoV-2 testing-related self-isolation using a generalized linear model to estimate the daily numbers of tests. The linear model employed detected SARS-CoV-2 cases and SARS-CoV-2 positivity rates from model output and historical Ontario data, respectively. (**Supplementary Text 2**). The ability to isolate while awaiting test results likewise depended on access to PSL and the capability to work remotely. We further assumed lost wages associated with severe COVID-19 regardless of access to PSL. Because of the uncertainty regarding the number of people unable to work due to severe COVID-19, we present results for a range of adherence-to-isolation scenarios.

Model parameters not informed by literature were calibrated using daily detected SARS-CoV2 cases as the calibration target under the base case scenario (**Supplementary Methods Text 1**). Each of the three PSL scenarios were simulated 100 times and summarized with means and credible intervals (CrI) generated from 0.025 and 0.975 quantiles. ABM analyses employed TreeAge (version 2021 R1.1, Williamstown, MA) and model outputs were compiled using R.^10–13^

## Results

In the simulated population, individuals with SARS-CoV-2 infection were younger, belonged to larger households, were more likely to be employed, and were more likely to be an essential worker (**Table1**). The estimated average number of cumulative, detected cases was 300,995 (95% CrI 279,038; 320,089) compared to the observed value of 303,745 (**Figure 1**).

**Table 1:** Characteristics of synthetic population and individuals infected with SARS-CoV-2 under 3 scenarios.

| Variable | Synthetic Population<sup>a</sup> | Base case mean (95% CrI) <sup>a</sup> | 3 Day PSL mean (95% CrI) <sup>a</sup> | 10 day PSL mean (95% CrI) <sup>a</sup> |
| --- | --- | --- | --- | --- |
| N<sup>a</sup> | 14,500,000 | 883,801 (822,434; 938,999) | 798,270 (713,205; 876,588) | 668,498 (471,763; 786,991) |
| Age, mean | 41.54 | 37.37 (37.19; 37.55) | 37.31 (37.09; 37.52) | 37.13 (36.77; 37.41) |
| Age, median | 41 | 37.07 (37; 38) | 37.04 (37; 38) | 36.94 (36; 37) |
| Age, 25<sup>th</sup> quantile | 23 | 20 (20; 20) | 19.99 (20; 20) | 19.62 (19; 20) |
| Age, 75<sup>th</sup> quantile | 60 | 53.97 (53.48; 54) | 53.96 (53; 54) | 53.69 (53; 54) |
| Female, % | 50.7% | 49.7% (49.4%; 50%) | 49.7% (49.4%; 50.1%) | 49.7% (49.4%; 50.1%) |
| Household size, mean | 3.14 | 4.13 (4.11; 4.15) | 4.14 (4.12; 4.16) | 4.16 (4.13; 4.21) |
| Employed FT, <sup>b</sup> % | 51.7% | 59.3% (59%; 59.7%) | 59.2% (58.8%; 59.6%) | 58.8% (57.8%; 59.4%) |
| Employed PT, % | 4.1% | 4.7% (4.6%; 4.9%) | 4.7% (4.5%; 4.9%) | 4.7% (4.5%; 4.9%) |
| Unemployed, % | 44.2% | 36% (35.6%; 36.3%) | 36.1% (35.7%; 36.6%) | 36.6% (35.9%; 37.6%) |
| Access to PSL, %<sup>b</sup> | 34.5% | 35.2% (34.8%; 35.6%) | 35.3% (34.8%; 35.8%) | 35.3% (34.7%; 35.9%) |
| Essential worker, % <sup>c</sup> | 66.6% | 74.5% (74.1%; 74.9%) | 74.4% (74%; 74.9%) | 74.1% (73.2%; 74.6%) |

<sup>a</sup>Counts from the 1.0 M synthetic population and agent based model were re-scaled upward by a factor of 14.5 to represent values on the Ontario provincial scale (14.5 M).
<sup>b</sup> Infected individuals in the model are more likely to belong to larger households, be in the labour force and be essential workers.
<sup>c</sup>Percentages are calculated among individuals in the labor force (workers).

**Figure 1:**
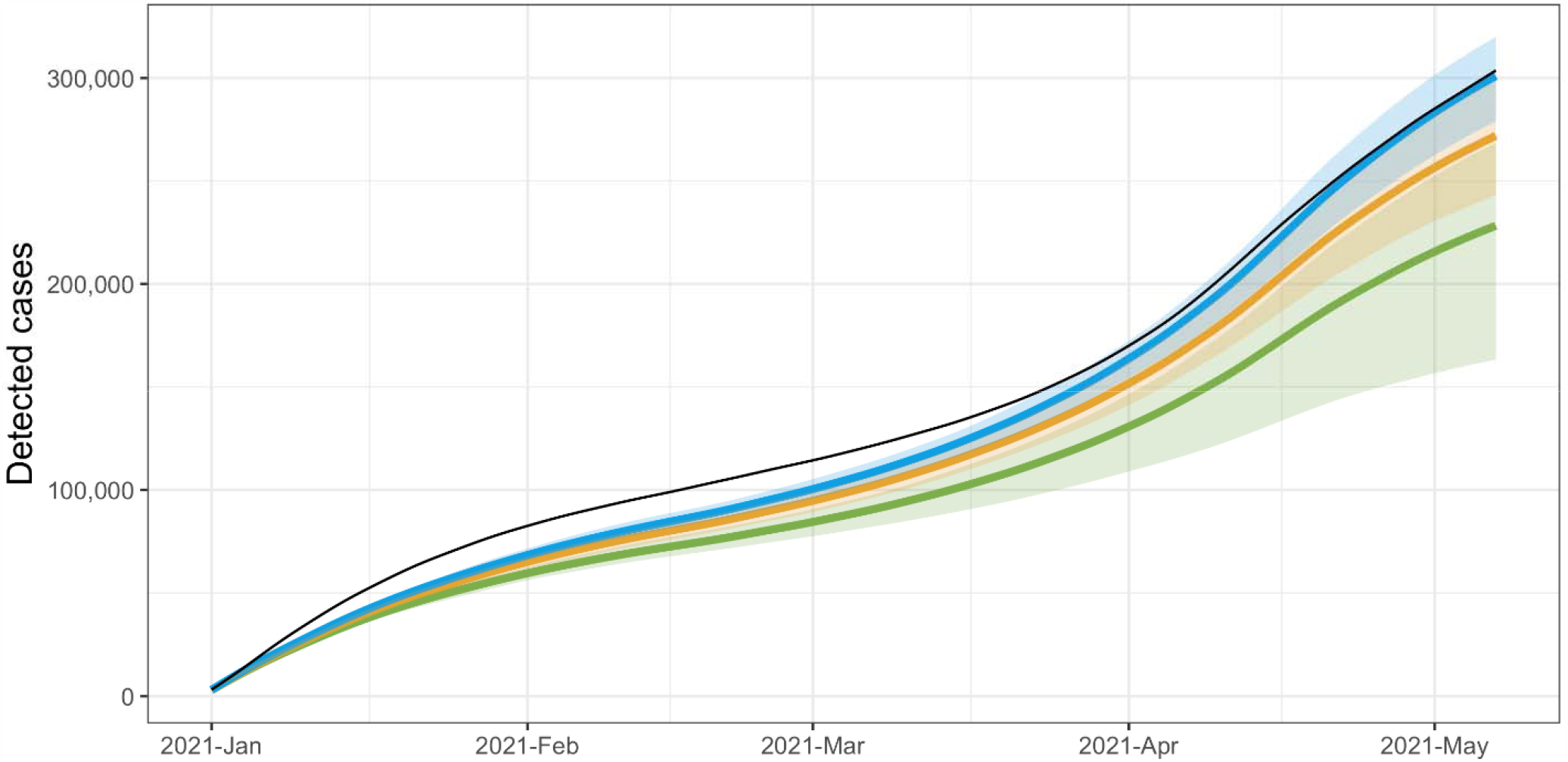
Cumulative number of detected SARS-CoV-2 cases from January 1, 2020, until May 1, 2020, in Ontario, Canada: observed (black), modelled with the base-case paid sick leave (PSL) policy (employer-provided only, blue), modelled with the 3-day universal PSL policy (employer-provided plus three PSL days for all remaining workers, orange), and modelled with a 10-day universal PSL policy (employer-provided plus ten PSL days for all remaining workers, green). 95% credible intervals are represented by the shaded areas.

### Health outcomes

In the base case, we estimated 883,800 (822,433; 938,998) cumulative infections during the study period, including both detected and undetected cases **(Supplementary Figure 1)**. In the 3- and 10-day universal PSL scenarios we estimated reductions in cumulative infections of 85,531 (-2,484; 195,318) and 215,302 (81,500; 413,742) associated with an increase of 7.9% (7%; 9.1%) and 9.5% (8%; 12.7%) of infected individuals self-isolating and 1,307 (-201; 3,205) and 3,352 (1,223; 6,528) fewer COVID-19 admissions, respectively, relative to the base case **(Table 2)**.

**Table 2:**
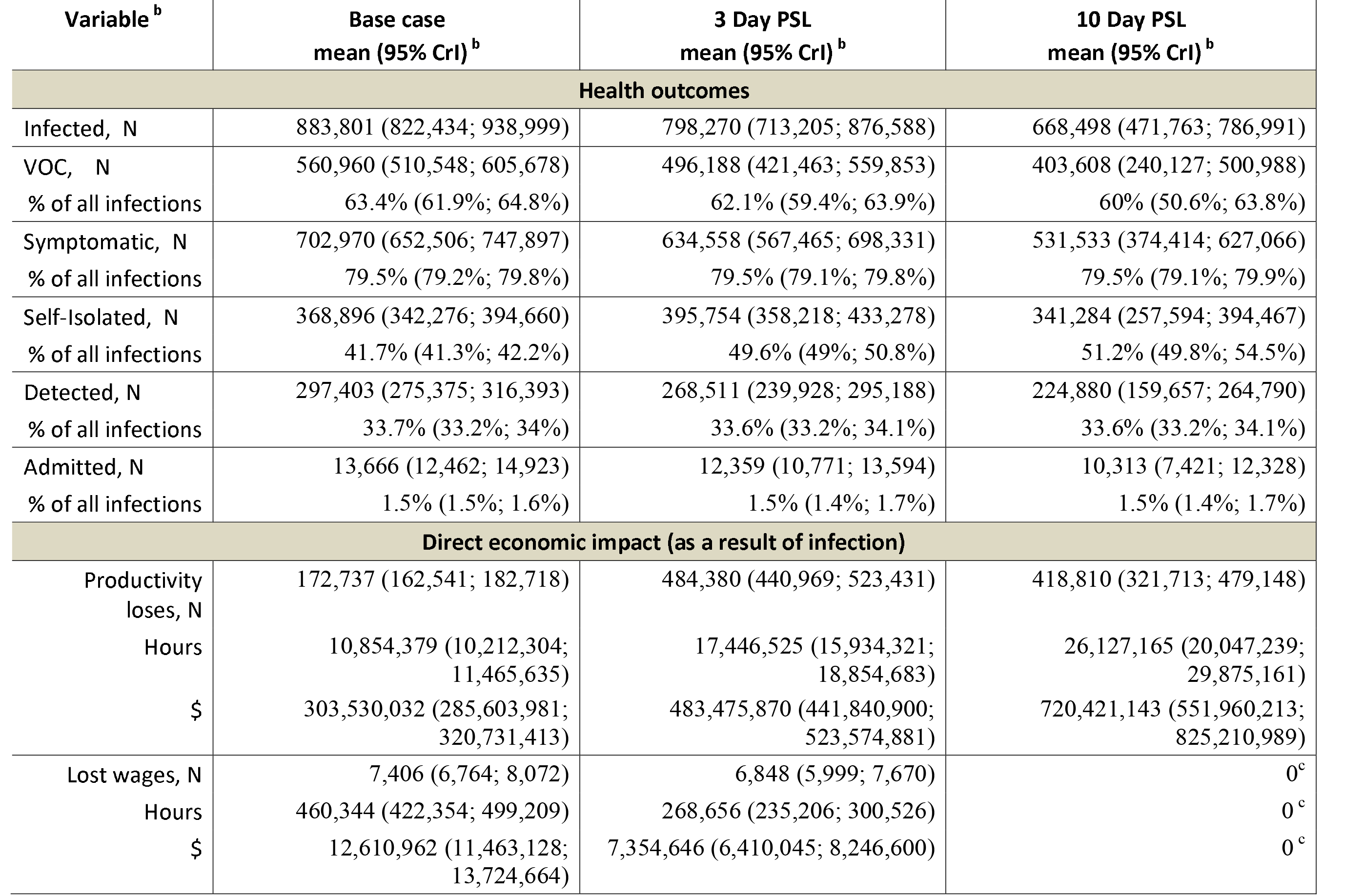
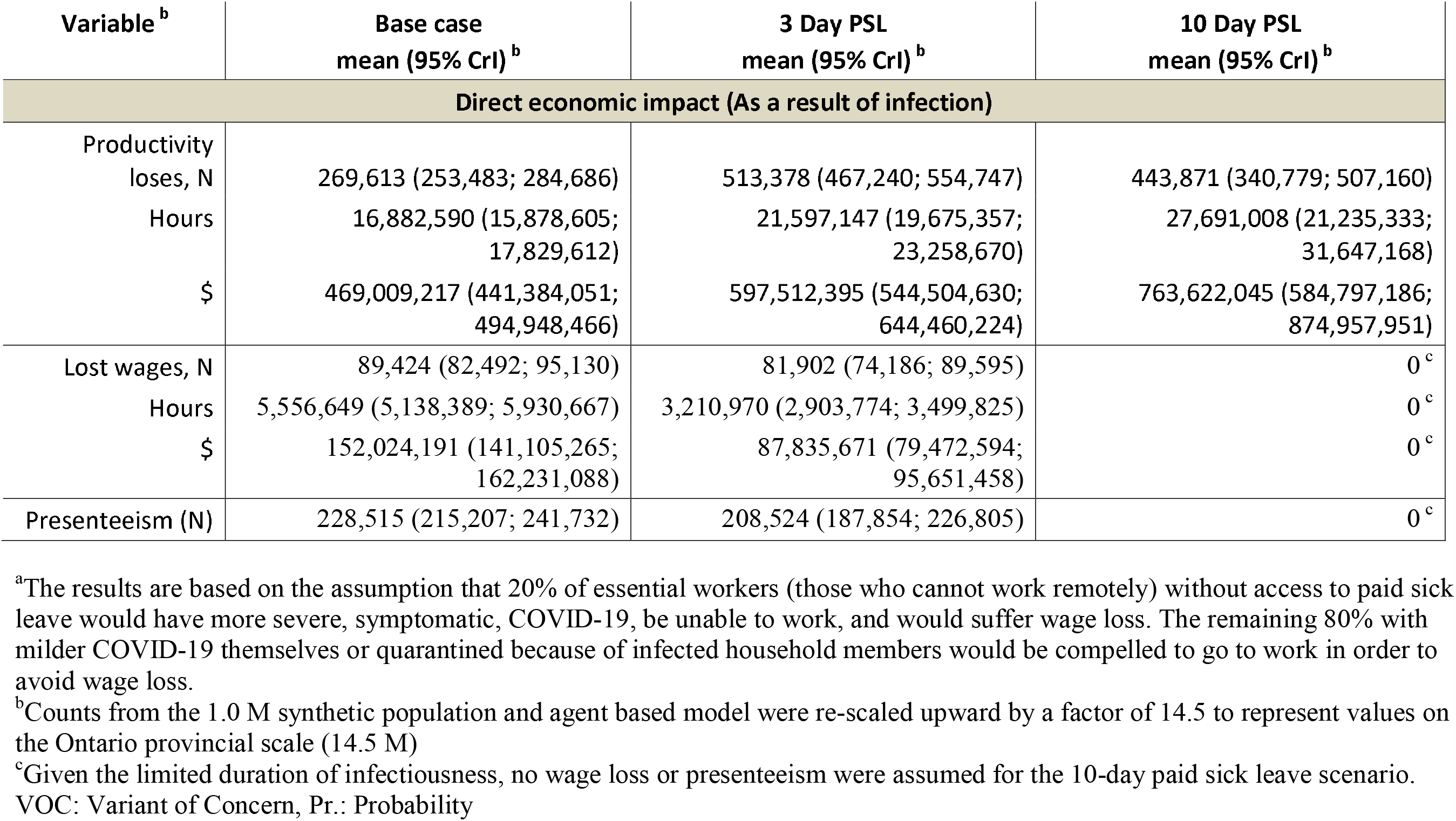
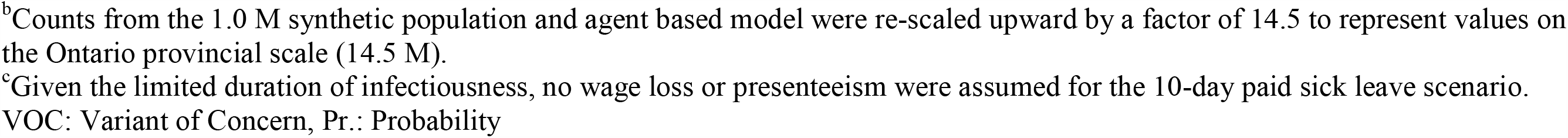
Health and Economic outcomes (1.5% scenario ^a^)

**Table 2a:**
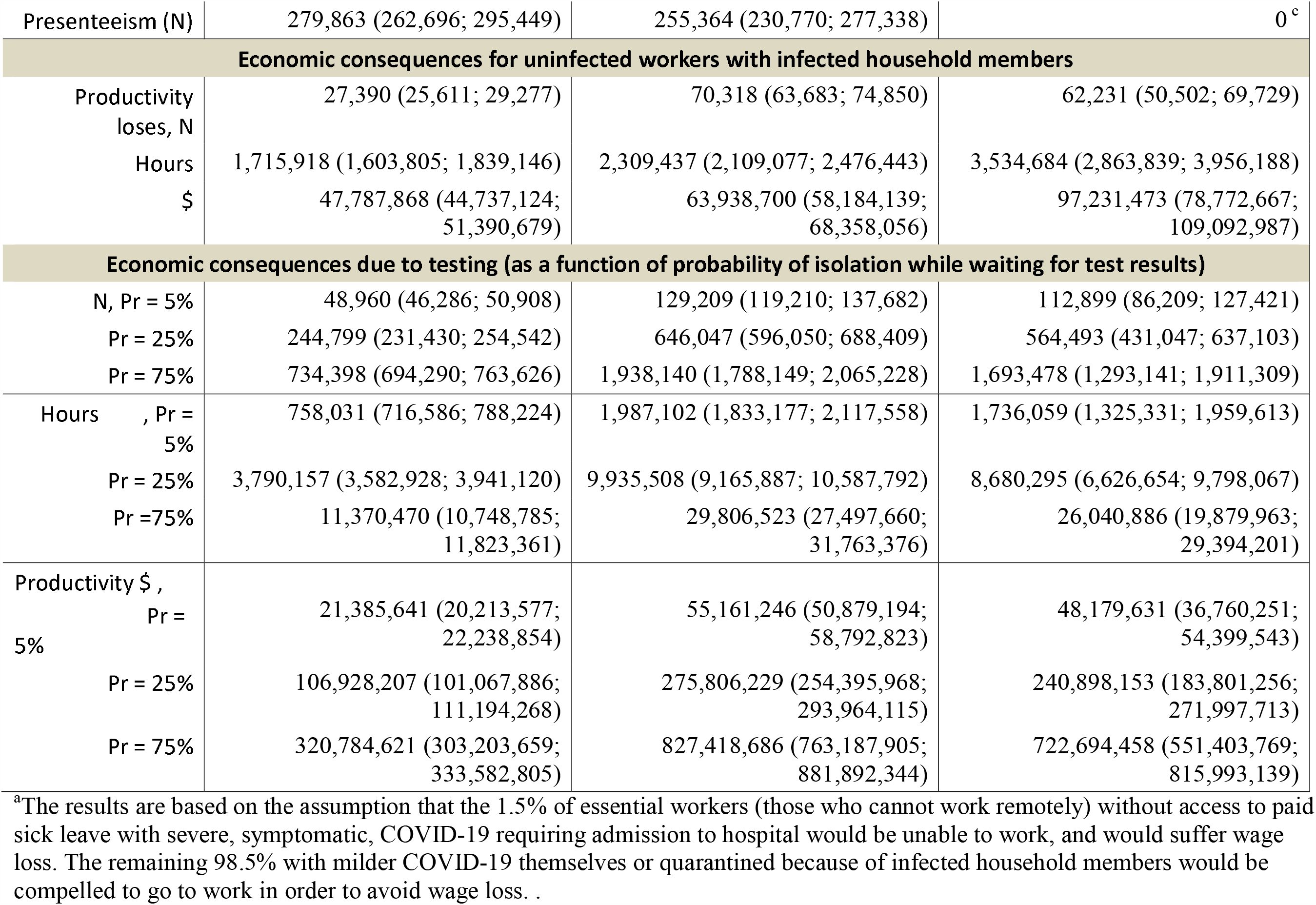
Health and Economic outcomes (20% scenario ^a^)

### Economic outcomes relating directly to SARS-CoV-2 infection

The estimated hours of productivity loss due directly to a SARS-CoV2 infection were 10,854,379 (10,212,304; 11,465,635) in the base case, 17,446,525 (15,934,321; 18,854,683) in the 3-day scenario, and 26,127,165 (20,047,239; 29,875,161) in the 10-day scenario. Fewer workers were estimated to engage in presenteeism, 24,499 (216; 54,170) and 279,863 (262,696; 295,449) and forego wages, 558 (-327; 1,608) and 7,406 (6,764; 8,072) in the 3- and 10-day scenarios compared to base case, respectively **(Table 2)**. Lost wages were $5,256,316 (4,077,280; 6,804,983) and $12,610,962 ($11,463,128; $13,724,664) lower in the 3 day and 10 day scenarios respectively, relative to the base case. Scenario analyses assuming increased proportion of severe disease resulted in larger estimates of lost-wages in the base case scenario compared to the 3 day and 10 day scenarios (**Table 2a**).

### Economic outcomes due to household member isolation & testing

Characteristics for household members of infected individuals are presented in **Supplementary Table 1**. Increases in hours of productivity loss among non-infected workers with infected household members were 593,519 (348,370, 804,471) in the 3-day and 1,818,766 (1,127,684; 2,317,681) in the 10-day PSL scenarios, relative to base case (**Table 2**). Similarly, productivity losses among workers isolating while awaiting test results ranged from 17,821,518 (15,332,584; 19,682,621) to 267,322,764 (229,988,756; 295,239,309 hours in the 3-day and 14,181,402 (8,454,490;17,388,008;) to 212,721,027 (126,817,350; 260,820,120) hours in the 10-day PSL scenarios, relative to base case, as adherence to self-isolation guidelines increased from 5 to 75% (**Table 2**).

## Discussion

We estimated that universal PSL policies reduce numbers of SARS-CoV-2 infections and COVID-19 hospitalizations. A 3-day universal PSL policy was estimated to produce a modest reduction in these outcomes whereas a 10-day policy produced a much larger reduction. An even longer PSL duration would be unlikely to reduce transmission further because infectivity with SARS-CoV-2 declines substantially after 8-10 days of infection. ^14^ For workers, universal PSL policies were estimated to reduce lost wages and presenteeism. From industry’s perspective, reduced presenteeism had opposing effects on worker productivity: indirectly protecting it due to fewer workers becoming infected but directly reducing it from workhours lost to isolation.

Our study has limitations: reduced long-term post-COVID-related disability, deaths prevented or quality-adjusted life years gained were not assessed; we assumed that workers would not forego wages in order to self-isolate except for severe COVID-19 disease; reduced unemployment or other increases in economic activity due to a reduction cases were not explicitly modeled; and the probability of employer-based PSL was obtained from 2016 census data. Further, we did not consider the effect of PSL to enable earlier diagnosis of SARS-CoV-2.

Further research could include cost-utility analyses and studies focussed on macroeconomic outcomes.

## Supporting information

Suppelmentary Material

## Data Availability

All data used in the present study originate from pblished literature and publically available sources.

## Acknowledgements

Not applicable

## Notes

### Competing Interest Statement

The authors have declared no competing interest.

### Author Declarations

All data used in the present study originate from pblished literature and publically available sources. references are provided in the main manuscript and in supplementary materials

