## Supplementary material for "Health and Economic Consequences of Universal Paid Sick Leave Policies During the COVID-19 Pandemic": Suppelmentary Material

**Supplementary Table 1** Characteristics of household members of infected individuals

| **Variable** | **Synthetic cohort^a^** | **Base Case^a^** | **10day^a^** | **3day^a^** |
| --- | --- | --- | --- | --- |
| N^a^ | 14,500,000 | 434,391 | 346,646 | 410,790 |
| Age, mean | 41.54 | 34.89 | 34.04 | 34.39 |
| median | 41 | 35.52 | 33.73 | 34.35 |
| 25^th^ Quantile | 23 | 18.03 | 16.8 | 17.08 |
| 75^th^ Quantile | 60 | 50.91 | 50.43 | 50.94 |
| % Female | 50.7% | 49.5% | 49.3% | 49.3% |
| Family Size, mean | 3.14 | 4.63 | 4.64 | 4.62 |
| Unemployed | 44.2% | 34.3% | 38.2% | 36.9% |
| Employed FT | 51.7% | 60.8% | 57.2% | 58.4% |
| Employed PT | 4.1% | 4.9% | 4.6% | 4.7% |

^a^Counts from the 1.0 M synthetic population and agent based model were re-scaled upward by a factor of 14.5 to represent values on the Ontario provincial scale (14.5 M).

**Supplementary Figure 1**

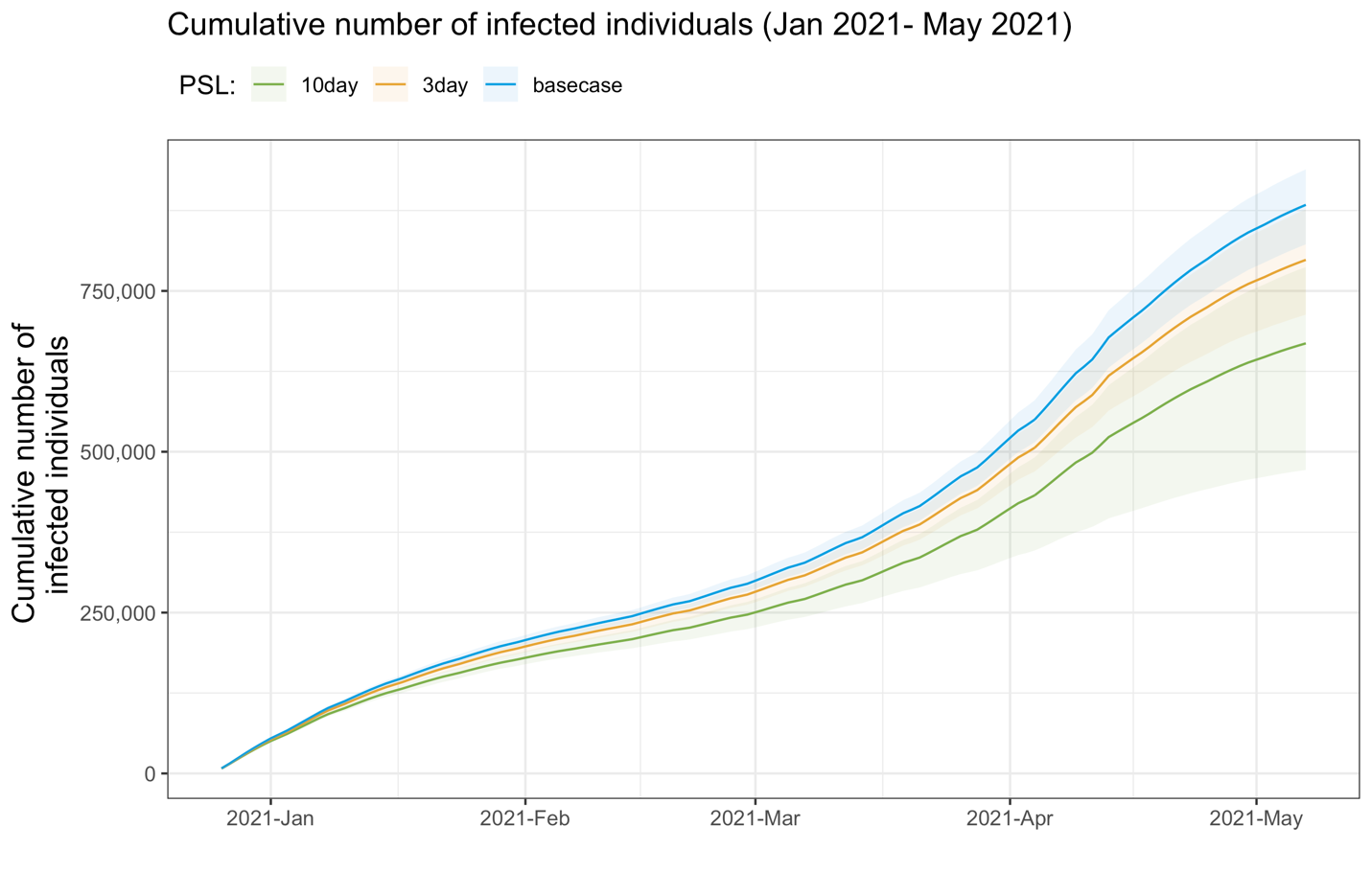

**Supplementary Figure 2**

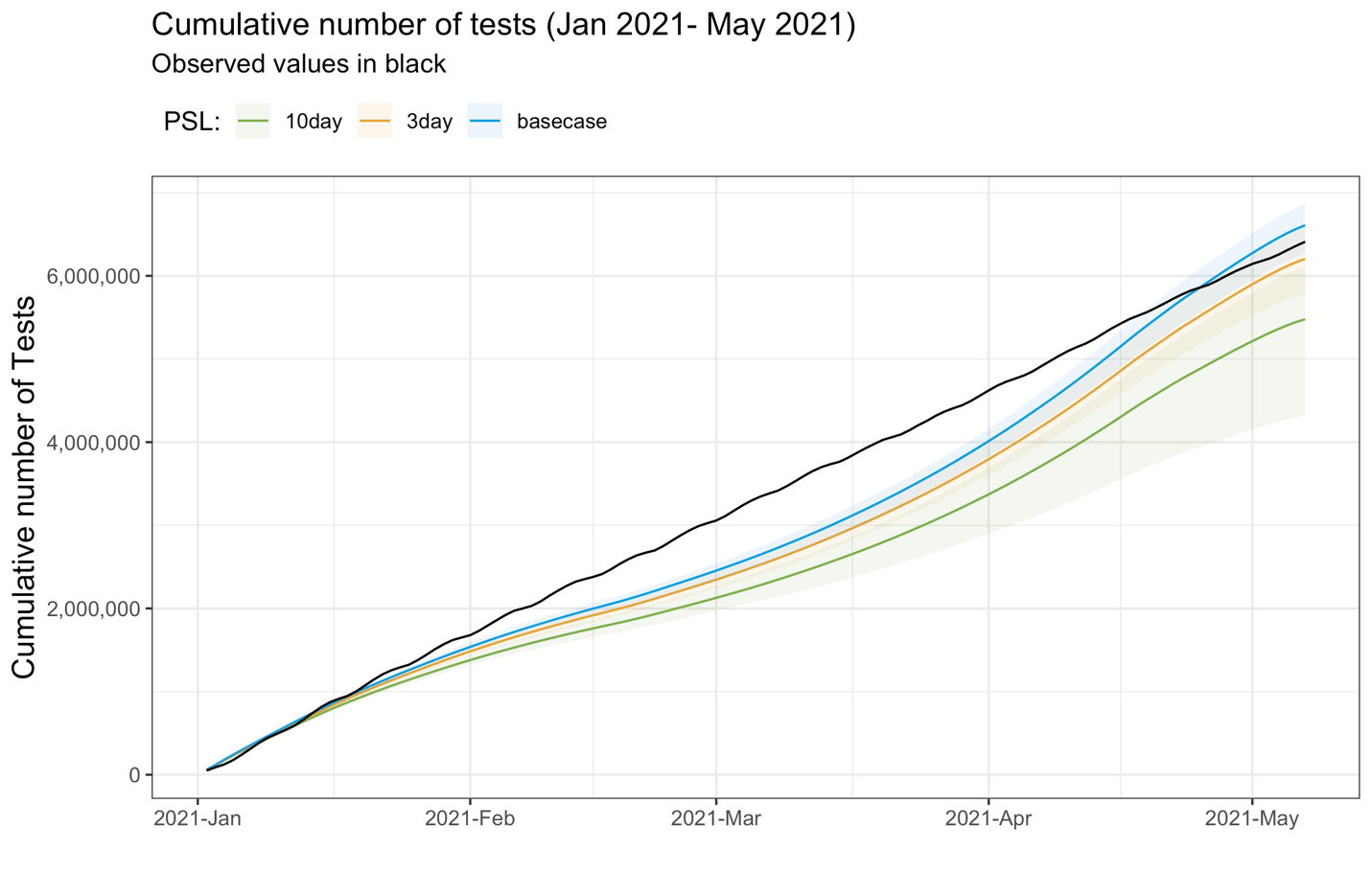

**Supplementary Text 1**

### Development of a synthetic population representative of Ontario, Canada

We developed a synthetic population of one million individuals representative of the residents of Ontario, Canada (population ~ 14.5 million) in 2020, based on data within the Social Policy Simulation Database and Model (SPSD/M) developed by Statistics Canada.^1^ SPSD/M contains data on Ontario households categorized by rural or urban location, numbers of household members, age and gender of household members, labour force participation and industry. For the synthetic population, households were randomly sampled from the available household types to yield a total of one million hypothetical individuals. Households were then randomly assigned to cities or a rural region according to the SPSD/M urban/rural designation of the associated household type. Within urban settings, households were further randomly assigned to neighborhoods while rural household were assigned to districts.

All members of the synthetic population spent time each day in their households, neighborhoods or districts, and their city or rural region. Additionally, children between 2 and 17 years old were assigned to educational settings (daycare, primary/elementary schools and high schools). Adults 18 to 34 years old could be enrolled in a post-secondary institution but, in analyses conducted to date, they were modelled to take classes remotely from their households. Adults 18 to 34 years old, not enrolled in a post-secondary institution, and those aged 35 to 64 years old could be members of the workforce. Workplaces were characterized by industry type, region, and workplace size (1 to 20, 21 to 99, 100 to 499 and 500 or more workers). Working age adults were then randomly assigned to workplaces in their region according to their SPSD/M industry designator. Adults, 65 and older, were assumed to be retired from the workforce.

### Agent-based model

Our agent-based model (ABM) for SARS-CoV-2 transmission made use of the synthetic population described above and was initially developed to represent the COVID-19 first and second waves in Ontario. During the latter, the ABM was used to estimate the change in incident and cumulative detected SARS-CoV2 infections associated with school opening on September 15, 2020 until October 31, 2020 compared to the counterfactual scenario in which schools had been kept closed.^2^ Details regarding the ABM methodology can be found in the online supplement of the school opening article.^2^ For the current analysis, the ABM was modified to represent the four-month period from January 1 to May 1, 2021, which constituted the COVID-19 third wave in Ontario (see the table below). The model randomly removed 40,288 individuals from the synthetic population (equivalent to 584,000 on the provincial scale) who had been infected with SARS-CoV-2 prior to the study period according to the distribution of the proportion of cases by ten-year age groups observed in Ontario. The ABM was then seeded with 8000 infectious individuals (equivalent to 116,000 individuals on the provincial scale) who were selected from the remaining synthetic population at random.

All other individuals, neither removed nor seed cases, were initially susceptible to SARS-CoV2 infection and could contact and be exposed to infectious individuals within their household, at school, at workplaces, in neighborhoods and cities, or rural districts. Exposed individuals may not have been infected and remain susceptible or they may have become infected. Infected individuals were unable to transmit the virus during a four-day latent period, after which, infected individuals became infectious and able to transmit the virus. Infectious individuals may or may not have developed symptoms. The former entered a one-day pre-symptomatic stage followed by a symptomatic stage. The duration of infectiousness for both individuals with and without symptoms was modelled to be 15 days.

A proportion of asymptomatic cases were modelled to be detected through contact tracing or other testing, reported as confirmed cases, and ordered to quarantine until recovery. Symptomatic infected individuals could seek health care, be confirmed as cases, and either be admitted to hospital or sent home and ordered to quarantine until recovered. Symptomatic individuals who did not seek health care had the possibility of self-isolating until recovery. Quarantined or self-isolated individuals could still transmit SARS-CoV2 to their household members. Uninfected workers could be ordered into quarantine by virtue of being in a household with an infected individual.

In the current analysis, the ABM was modified such that the ability of infected workers to adhere to a quarantine order or to self-isolate was dependent on type of work (essential or non-essential) and access to paid sick leave (PSL) whether provided by the employer or as part of a 3- or 10-day universal PSL policy. Essential work was defined as a type of employment that could not be performed remotely and required the worker to be present at a workplace. The ABM assumed that essential workers without any form of PSL would be compelled to go to their workplace (i.e. to engage in presenteeism) to avoid wage loss unless they suffered from severe COVID-19. Data regarding the proportion of essential workers without PSL who would stay away from their workplace is lacking. Two scenarios were therefore modelled: a conservative scenario where only workers ill enough to be admitted to hospital would avoid their workplace and a more liberal scenario in which 20% of workers would stay home despite wage loss. Likewise, data is lacking on the probability that an uninfected essential worker without PSL would adhere to a quarantine order due to an infected household member. We estimated the economic consequences of PSL policy assuming a 5, 25 and 75% compliance with quarantine orders.

*Agent-based model (ABM) inputs and calibration*

|  | **Value** | **Source** |
| --- | --- | --- |
| **SARS-CoV2 epidemiology** | | |
| Latent period | 4 days | Bi^3^, Gostic^4^ |
| Incubation period | 5 days | Kucharski^5^ |
| Infectious period | 15 days | Voinski^6^ |
| **Daily contact numbers** | | |
| Household (n, range) | 1 - 7 | SPSD/M database^1^ |
| Neighborhood/district^a^ (mean, SD) | 1.5 (1.1) | CONNECT study^7^ |
| Region^a^ (mean, SD) | 1.8 (1.2) | CONNECT study^7^ |
| School^a^ (mean, SD) | 5 (1.4) | CONNECT study^7^ |
| Work^a^ (mean, SD) | 10 (1.6) | CONNECT study^7^ |
| College or university campus | 15 | Calibrated value^b^ |
| Proportionate reduction in non-household contacts Jan. 1 to Feb. 13, 2021, compared to pre-pandemic | 0.45 | Calibrated value^b^ |
| Proportionate reduction in non-household contacts Feb. 14 to Apr. 14, 2021, compared to pre-pandemic | 0.7 | Calibrated value^b^ |
| Proportionate reduction in non-household contacts Apr. 15 to May 1, 2021, compared to pre-pandemic | 0.81 | Calibrated value^b^ |
| **Transmission probability per contact** | | |
| Household – children <= 10 years old | 0.024 | Calibrated value^b^ |
| Household – children > 10 years old | 0.116 | Li^8^, Kucharski^5^, Bi^3^, Cheng^9^. Refined via calibration^b^ |
| Household – adults | 0.115 | Kucharski^5^, Bi^3^, Cheng^9^. Refined via calibration^b^ |
| Other settings | 0.033 | Kucharski^5^, Bi^3^, Cheng^9^. Refined via calibration^b^ |
| Proportionate reduction due to asymptomatic status | 0.90 | Kucharski^5^. Refined via calibration^b^ |
| **Case detection probability** | | |
| Symptomatic | 0.4 | Tuite^10^, PHO^11^. Refined via calibration^b^ |
| Asymptomatic | 0.3 | Tuite^10^, PHO^11^. Refined via calibration^b^ |

SD – standard deviation, PHO – Public Health Ontario

^a^Standard deviation of the log contact number assumed to be 0.2*mean log contact number with an assumed correlation coefficient among settings of 0.9.

^b^Calibration refers to adjustment of the model input values derived from jurisdictions other than Ontario or determining the most likely input value for parameters unavailable from data or literature sources to minimize the difference between modelled and observed daily, new, confirmed SARS-CoV2 cases in the third wave of the pandemic in Ontario from January 1 to May 1, 2021.

**Supplementary Text 2**

*PSL Estimation*

The probability of having paid sick leave was estimated from Statistics Canada’s General Society Survey (GSS), Cycle 30: Canadians at Work and Home.^12^ The GSS has a targeted sample size of approximately 20,000 Canadians over the age of 15, with questionnaires being administered online and via telephones. A comprehensive description of the survey design, data collection and methodology can be found on the Statistics Canada website. Cycle 30 of the GSS collected information on lifestyle and workplace information during 2016. We used a logistic regression for complex survey designs and included age, sex, marital Status, Income, and family income as covariates.

*Estimating COVID-19 tests from Cases*

We sourced testing and cases stratified by age from the Ontario Data Catalogue for the study period. We then fit 6 linear models predicting the percent positive on a specific day as a function of total positive Covid-19 cases on that day. We estimate the linear model for each of the age categories (0 to 13,14 to 17, 18 to 24, 25 to 64, and 65+) available in the Ontario Data Catalogue. We than used the daily predicted detected infections from the ABM and the coefficients for the linear model to predict the total number of COVID-19 tests on each day in the study period.

**Supplementary Text 3**

*Assignment of infected individuals’ productivity losses*

The following table indicates the calculated productivity loss in days for workers infected with SARS-CoV2 and in the labor force assuming that 20% of essential workers without PSL would stay home despite suffering wage losses.

| **Symptomatic** | **Essential Worker** | **Chose to Self isolate** | **Detected overall (Via testing)** | **Gets PSL** | **Productivity impact (days)** | | |
| --- | --- | --- | --- | --- | --- | --- | --- |
|  |  |  |  |  | Base case | 3 day PSL | 10 day PSL |
| No | No | No | No | No | 0 | 0 | 0 |
| No | No | No | No | Yes | 0 | 0 | 0 |
| No | No | No | Yes | No | 0 | 0 | 0 |
| No | No | No | Yes | Yes | 0 | 0 | 0 |
| No | Yes | No | No | No | 0 | 0 | 0 |
| No | Yes | No | No | Yes | 0 | 0 | 0 |
| No | Yes | No | Yes | No | 0 | 3 | 10 |
| No | Yes | No | Yes | Yes | 11.6 ^a^ | 11.6 | 11.6 |
| Yes | No | No | No | No | 2.32 ^b^ | 2.32 | 2.32 |
| Yes | No | No | No | Yes | 2.32 | 2.32 | 2.32 |
| Yes | No | No | Yes | No | 2.32 | 3 | 10 |
| Yes | No | No | Yes | Yes | 11.6 | 11.6 | 11.6 |
| Yes | No | Yes | No | No | 2.32 | 3 | 10 |
| Yes | No | Yes | No | Yes | 11.6 | 11.6 | 11.6 |
| Yes | No | Yes | Yes | No | 2.32 | 3 | 10 |
| Yes | No | Yes | Yes | Yes | 11.6 | 11.6 | 11.6 |
| Yes | Yes | No | No | No | 2.32 | 3 | 10 |
| Yes | Yes | No | No | Yes | 2.32 | 2.32 | 2.32 |
| Yes | Yes | No | Yes | No | 2.32 | 3 | 10 |
| Yes | Yes | No | Yes | Yes | 11.6 | 11.6 | 11.6 |
| Yes | Yes | Yes | No | No | 2.32 | 3 | 10 |
| Yes | Yes | Yes | No | Yes | 11.6 | 11.6 | 11.6 |
| Yes | Yes | Yes | Yes | No | 2.32 | 3 | 10 |
| Yes | Yes | Yes | Yes | Yes | 11.6 | 11.6 | 11.6 |

^a^With a five-day workweek, a 15-day infectious period equates to 11.6 workdays,

^b^2.32 = 20% of symptomatic individuals with severe disease *11.6

*Assignment of productivity losses for uninfected workers with infected household members*

| **FT/PT** | **Essential** | **Gets PSL** | **Productivity impact (days)** | | |
| --- | --- | --- | --- | --- | --- |
|  |  |  | Base case | 3 day PSL | 10 day PSL |
| FT | No | Yes | 0 | 0 | 0 |
| FT | No | No | 0 | 0 | 0 |
| FT | Yes | Yes | 11.6 | 11.6 | 11.6 |
| FT | Yes | No | 0 | 3 | 10 |
| PT | No | Yes | 0 | 0 | 0 |
| PT | No | No | 0 | 0 | 0 |
| PT | Yes | Yes | 11.6 | 11.6 | 11.6 |
| PT | Yes | No | 0 | 3 | 10 |

*Assignment of productivity losses due to testing*

| **Essential** | **Gets PSL** | **Productivity impact (days while waiting test results )** | | |
| --- | --- | --- | --- | --- |
|  |  | Base case | 3-day PSL | 10-day PSL |
| Yes | No | 0 | 2 | 2 |
| Yes | Yes | 2 | 2 | 2 |
| No | No | 0 | 0 | 0 |
| No | Yes | 0 | 0 | 0 |

11.	Ontario Ministry of Health. *Integrated Public Health Information Systems (IPHIS)*.; 2020.

12.	Statistics Canada. *The General Social Survey: Cycle 30 Public Use Microdata File*.; 2016. https://www150.statcan.gc.ca/n1/pub/89f0115x/89f0115x2019001-eng.htm
